# Knowledge syntheses in medical education: Examining author gender, geographic location, and institutional affiliation

**DOI:** 10.1101/2021.03.01.21252622

**Authors:** Lauren A. Maggio, Anton Ninkov, Joseph A. Costello, Erik W. Driessen, Anthony R. Artino

**Affiliations:** Professor of Medicine and the Associate Director of Scholarly Communication at Uniformed Services University of the Health Sciences in Bethesda, Maryland, USA. @LaurenMaggio; Postdoctoral Fellow at University of Ottawa, School of Information Studies in Ottawa, Ontario Canada. @TheNinkov; Research Assistant at the Henry Jackson Foundation and Uniformed Services University of the Health Sciences in Bethesda, Maryland, USA; Professor of Medical Education at Faculty of Health Medicine and Life Sciences, Maastricht University, Maastricht, The Netherlands. @erikwdriessen; Professor of Health and Human Function at the George Washington University School of Medicine and Health Sciences, Washington, DC, USA. @mededdoc

## Abstract

**Purpose:** Authors of knowledge syntheses make many subjective decisions during their review process. Those decisions, which are guided in part by author characteristics, can impact the conduct and conclusions of knowledge syntheses, which assimilate much of the evidence base in medical education. Therefore, to better understand the evidence base, this study describes the characteristics of knowledge synthesis authors, focusing on gender, geography, and institution.

**Method:** In 2020, the authors conducted a case study of authors of 963 knowledge syntheses published between 1999 and 2019 in 14 core medical education journals using a publicly accessible dataset.

**Results:** The authors of the present study identified 4,110 manuscript authors across all authorship positions. On average there were 4.3 authors per knowledge synthesis (SD=2.51, Median=4, Range=1-22); 79 knowledge syntheses (8%) were single-author publications. Over time, the average number of authors per synthesis increased (M=1.80 in 1999; M=5.34 in 2019). Knowledge syntheses were authored by slightly more females (n=2047; 50.5%) than males (n=2005; 49.5%) across all author positions (Pearson X^2^=22.02, *p*<.001). Authors listed affiliations in 58 countries, and 58 knowledge syntheses (6%) included authors from low- or middle-income countries (LMIC). Authors from the United States (n=366; 38%), Canada (n=233; 24%), and the United Kingdom (n=180; 19%) published the most knowledge syntheses. Authors listed affiliation at 617 unique institutions, and first authors represented 362 unique institutions with greatest representation from the University of Toronto (n=55, 6%) and the Mayo Clinic (n=31, 3%). Across all authorship positions, the large majority of knowledge syntheses (n=753; 78%) included authors at top 200 ranked institutions.

**Conclusions:** Knowledge synthesis author teams have grown over the past 20 years, and while there is near gender parity across all author positions, authorship has been dominated by North American researchers located at highly ranked institutions. This suggests a potential overrepresentation of certain authors with particular characteristics, which may impact the conduct and conclusions of knowledge syntheses in medical education.

## Introduction

In medical education, researchers have been encouraged to publish knowledge syntheses and educators to act as evidence-informed practitioners in their application of these reviews.^1,2^ As a result, the recent proliferation of knowledge syntheses published in core medical education journals is unsurprising.^3^ Knowledge syntheses, which often form the evidence base for implementing curricular innovations to determining how a field defines its key terminology, can have immense impact on a field’s discourse and future directions.^4,5^

When conducting a knowledge synthesis, author teams are required to make multiple decisions, many of which can be subjective. For example, authors must decide which populations to include and which to exclude; which contexts matter and which do not; which factors are important to extract from the primary studies and which are not; and even which languages to review and which to exclude. Such decisions are shaped by author characteristics, backgrounds, and even the power structures and cultural norms in which the authors operate. Therefore, best practices in knowledge syntheses encourage scholars to assemble a diverse author team with representation from a variety of backgrounds and perspectives.^6-8^ As such, these factors have the potential to impact the conduct of any given knowledge synthesis, as well as the conclusions drawn from the analysis.^9^ In short, author characteristics have important implications for a field’s evidence base.

Broadly speaking, researchers have raised concerns that author characteristics, such as gender,^10-13^ geographical location,^14-16^ and institutional affiliation^17,18^ can bias publications, including knowledge syntheses, and inadvertently reinforce dominant power structures. To this point, the Cochrane Collaboration, a major supporter and publisher of systematic reviews, has flagged the lack of international representation and diversity in published reviews as a significant problem and reports that more diverse author teams generate more relevant reviews with less research waste and fewer errors.^19^ In 2020, based in part on these findings, the Cochrane Collaboration advocated for wide participation from a variety of stakeholders in conducting reviews as one of its key strategic initiatives.^20^

In medical education, we know little about the characteristics of the authors who write knowledge syntheses, and so we lack a clear understanding of which author voices dominate and which are absent from the evidence base created through these reviews. This gap in our understanding means we risk inadvertently prioritizing certain views while diminishing others, and potentially creating an evidence base that is irrelevant to some people in some contexts. For example, a review written by a United States author team on providing student feedback may resonate well with North American readers.

However, due to a series of author decisions (e.g., inclusion criteria) and interpretations that likely vary based on cultural norms, such a review may be less useful to an audience outside of North America where cultural norms and educational systems differ.

Medical education researchers have just begun examining authorship characteristics. For example, a recent study explored author gender from articles published in four medical education journals,^21^ and another investigated authors’ geographic location in papers indexed as medical education.^22^ While both of these recent studies are valuable, they do not specifically examine knowledge syntheses nor do they examine the interplay of these factors.

In this study, we describe and examine the characteristics of the authors of knowledge syntheses with a focus on gender, geographical location, and institutional affiliation. In addition, where possible, we examine the interplay between these three factors. In doing so, we hope to raise medical educators’ awareness of author characteristics that may have bearing on the current state of the field’s evidence base. We also hope to foster a healthy skepticism in the evidence base that has been and continues to be created in medical education through knowledge syntheses.

## Methods

We conducted a case study of authors of knowledge syntheses published in 14 core medical education journals between 1999-2019.

### Data Collection

To undertake this case study, we utilized a publicly accessible data set that members of this author team created in 2020.^23^ The dataset includes citations and related PubMed metadata for 963 knowledge syntheses published in 14 core medical education journals between 2009-2019 (See Supplemental Appendix A for complete journal list). Full details of the creation of the data set are published in Maggio et al., 2020.^3^ We utilized this existing data set because data reuse has been associated with reduced research waste, faster translation of research findings into practice, and enhanced reproducibility and transparency of science.^24,25^ Specifically, we chose this data set because it is the only existing, up-to-date data set of knowledge syntheses in medical education. All data were downloaded and managed in GoogleSheets.^26^

### Data Enrichment

To determine author gender, we extracted the first names of all authors from the data set. In cases where authors used initials only (e.g., D.A.D.C Jaarsma, aka Debbie Jaarsma), we conducted a web search to identify their first name. All first names were then submitted to the gender prediction tool Genderize.io.^27^ Genderize.io predicts whether a name is male or female based on a database of over 20,000 names and provides a probability that the name is either male or female. This tool has been used in multiple publications with similar aims to the current study (e.g., Hart and Perlis^28^; Bagga et al.^29^). We accepted the tool’s designation for a name if the probability was over 70%. For each name that Genderize.io reported with <70% certainty (n=151), we looked up the authors’ online presence and cross referenced the authors’ names with their publication and online profiles at their academic institutions and social media sites (e.g., LinkedIn, Academia.edu, ResearchGate). In addition to the low certainty names, Genderize.io failed to identify 90 first names. For these names, we used the same web-based approach as described above for each author.

### Geographical Location

For each knowledge synthesis, we extracted from the PubMed metadata the country of all author institutions. Due to the structure of the metadata, we were only able to accurately identify the location of the first author at the level of the individual author. Therefore, for authors other than the first author we report for each knowledge synthesis all countries represented without regard to an individual’s placement in author order. Countries were described using the World Bank’s 2021 world region classification system,^30^ which includes four-levels of countries (low, lower middle, uppermiddle, and high income). Country levels are based on a country’s gross national income.^30^

### Institutions

For each knowledge synthesis, we identified the first author’s institutional affiliation. Similar to geographical location, we also identified for each knowledge synthesis a listing of all institutions that contributed to the study. To characterize institutional affiliation, we used the Times Higher Education (THE) World University Rankings for 2020.^31^ We selected this ranking due to its broad coverage of over 1,400 universities from 92 countries. THE ranking is based on 13 metrics (e.g., teaching, research, international outlook, industry outcomes). This ranking groups institutions starting with the top 200 institutions individually and then reports the remaining institutions in groups of 50, 100, or 200 depending on their position. In a minority of the cases examined in this study, authors provided multiple affiliations. In such cases, we included the first affiliation listed. Additionally, authors listed non-academic affiliations (e.g. professional associations, government entities, community hospitals), which we coded as organizations. Organizations are accounted for in our results, but unranked in relation to the THE rankings.

### Analysis

Descriptive and inferential statistics were calculated using GoogleSheets (Google, 2021), and data were visualized using Tableau v.2020.04.^32^

## Results

We identified 4,110 authors listed across all authorship positions, and of those 3,199 were unique authors (See Supplemental Appendix B for a listing of the most prolific authors). The number of authors per knowledge synthesis ranged from 1-22 with an average of 4.27 authors (SD=2.51, Median=4). Seventy-nine knowledge syntheses (8.2%) were single-author publications. Over the 20-year time period examined, the average number of authors per knowledge synthesis increased (M=1.80 in 1999; M=5.34 in 2019; See Figure 1).

**Figure 1:**
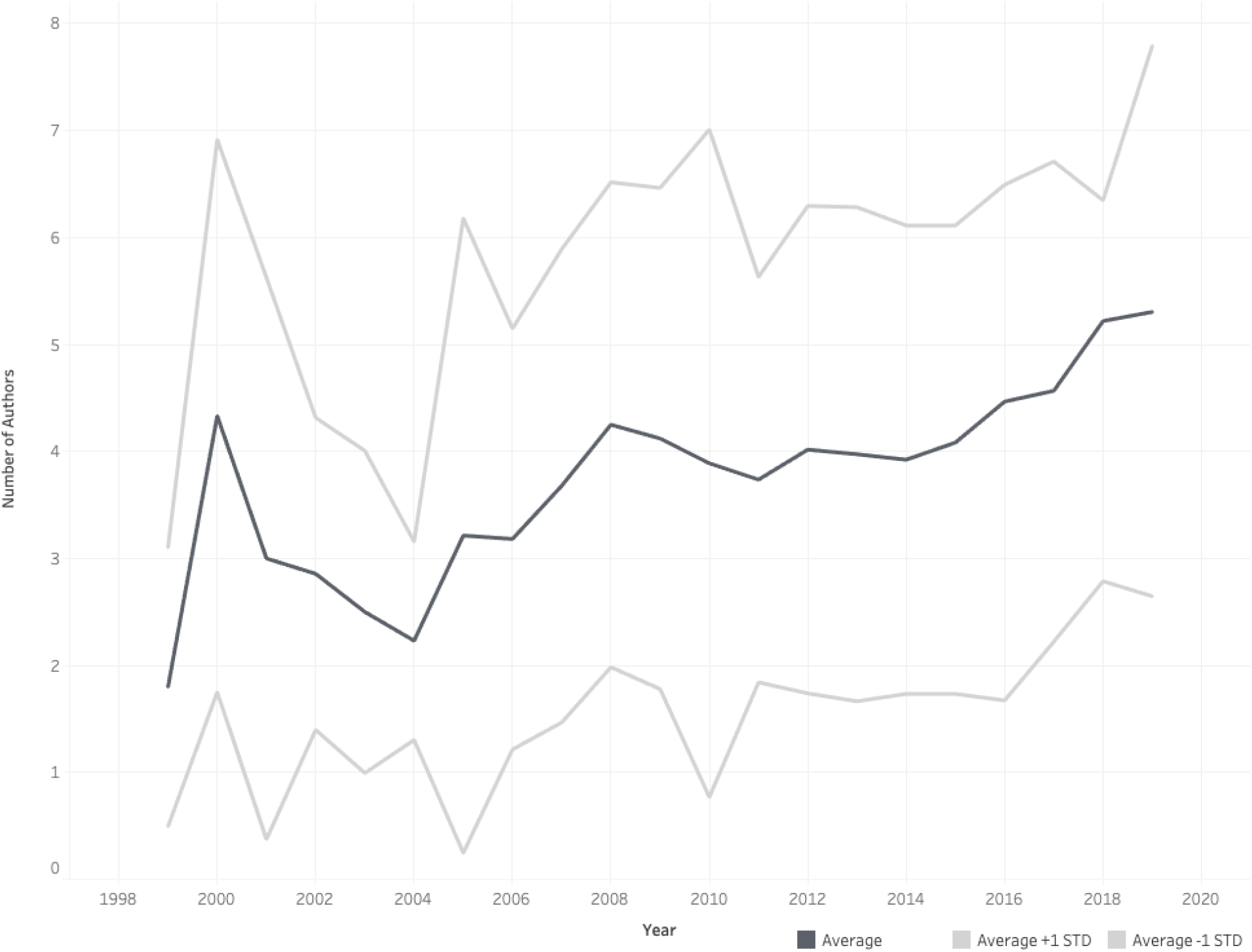
The average number of authors per knowledge synthesis (plus or minus 1 standard deviation) published in 14 core medical education journals published between 1999-2019

### Gender

We identified the gender for 4052 authors. We were unable to make a confident evaluation of gender identification for 58 authors (even after a web search), all of whom only appeared once in the dataset. Knowledge syntheses were authored by slightly more females (n=2047; 50.5%) than males (n=2005; 49.5%) across all author positions (Pearson X^2^=22.02, *p*<.001). In addition, more females were listed as first authors (n=494; 51.9%) and second authors (n=483; 55.4%), although only the differences in second author position were statistically significant (Pearson X^2^=8.49, *p*<.01). On the other hand, the last author position was held by more males (n=404; 56.0%), and this difference was statistically significant (Pearson X^2^=12.63, *p*<.001). Finally, there were no differences in the middle author position (Pearson X^22^=0.18, *p*=.67). See Figure 2 for author order by gender.

**Figure 2:**
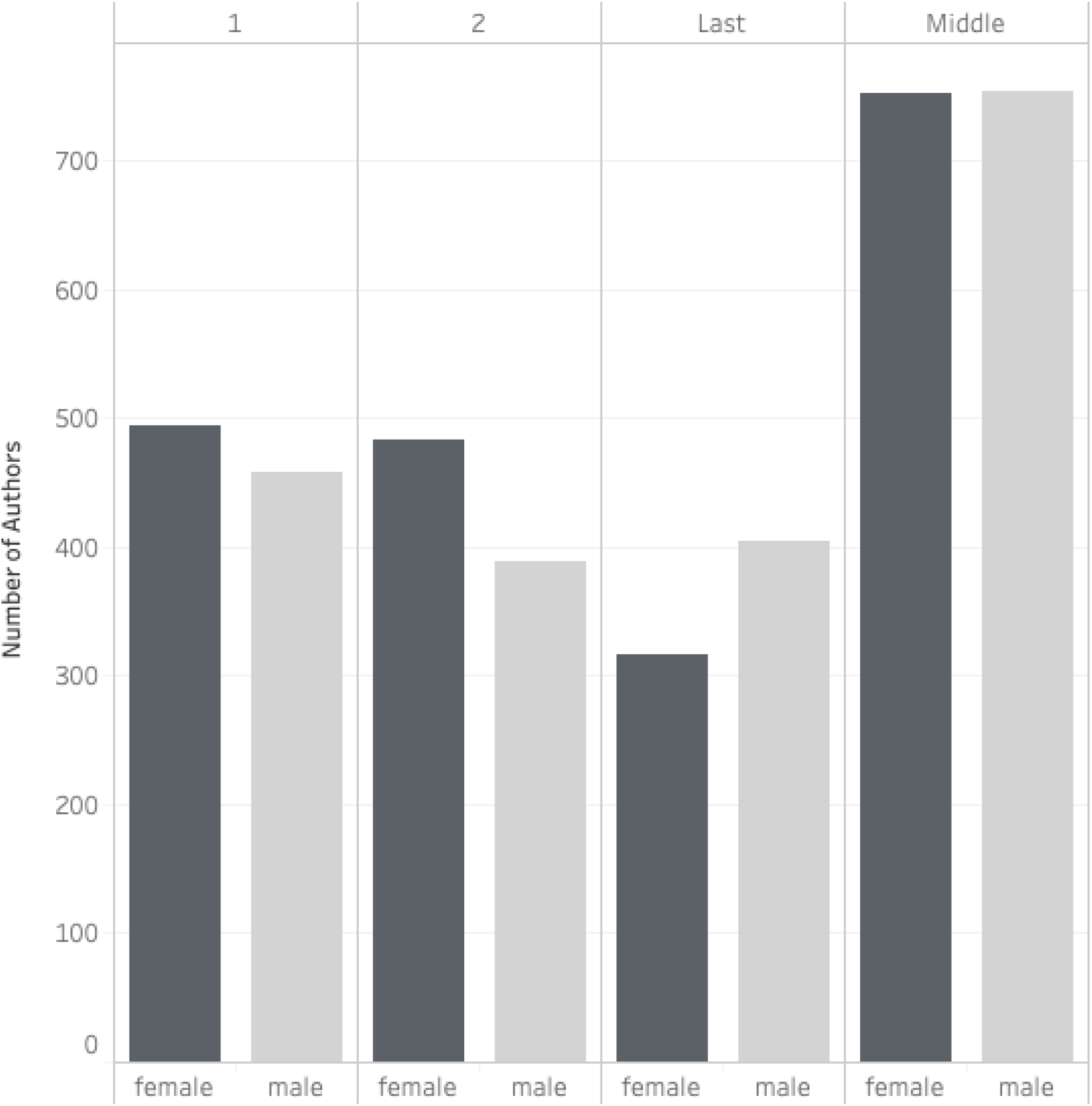
Author order by gender for knowledge syntheses in 14 core medical education journals published between 1999-2019. We were unable to determine the gender of 58 author names, which are excluded from this figure. *p<.01; **p<.001

Most author teams were a combination of genders (n=683 teams; 70.9%), but 280 teams included authors of a single gender (117 all female; 163 all male). For single authored papers, 52 were written by males and 26 by females. Over the time period examined, the ratio of female authors in all positions has increased (See Figure 3).

**Figure 3:**
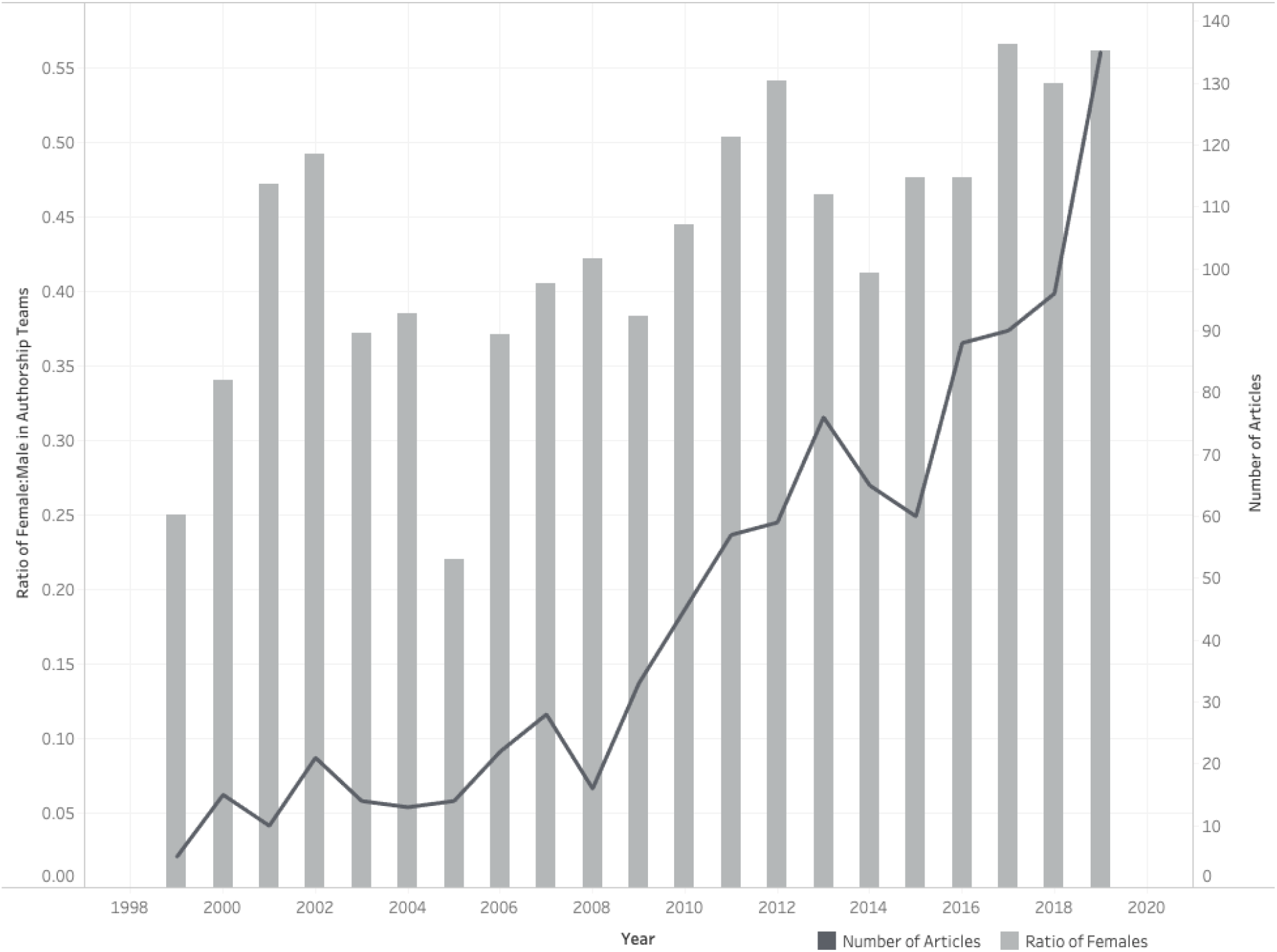
The ratio of female authors in all authorship positions for knowledge syntheses published in 14 core medical education journals published between 1999-2019.

### Geography

Across all authorship positions, authors listed affiliations in 58 countries, including 22 LMIC (see Figure 4; See Supplemental Appendix C for listing of all countries). By number of knowledge syntheses, the countries most represented were the United States (US) (n=366; 38%), Canada (n=233; 24%), and the United Kingdom (UK) (n=180; 19%). Fifty-eight (6%) knowledge syntheses included at least one author listing an affiliation based in a LMIC; of these, 39 (4%) were first authored by an author with a LMIC affiliation. Of the 58 knowledge syntheses including LMIC authors, authors based in China were most prolific, publishing 22 knowledge syntheses of which 15 were written by authors all based in China. First authors represented 42 countries, including 13 LMIC. The most represented countries for first authors were the US (n=312; 25%), Canada (n=183; 15%), and the UK (n=151; 12%). Twenty-seven (3%) knowledge syntheses were exclusively authored by individuals based in a LMIC. The most countries represented on a single team were seven, in a study featuring authors from France, Ireland, UK, Italy, Belgium, Croatia, and Germany.^33^ Eighty percent (n=767) of knowledge syntheses included authors from a single country only. Of those representing a single country, authors were predominantly located in the US (n=271; 22%), Canada (n=149; 12%), and the UK (n=122; 10%).

**Figure 4:**
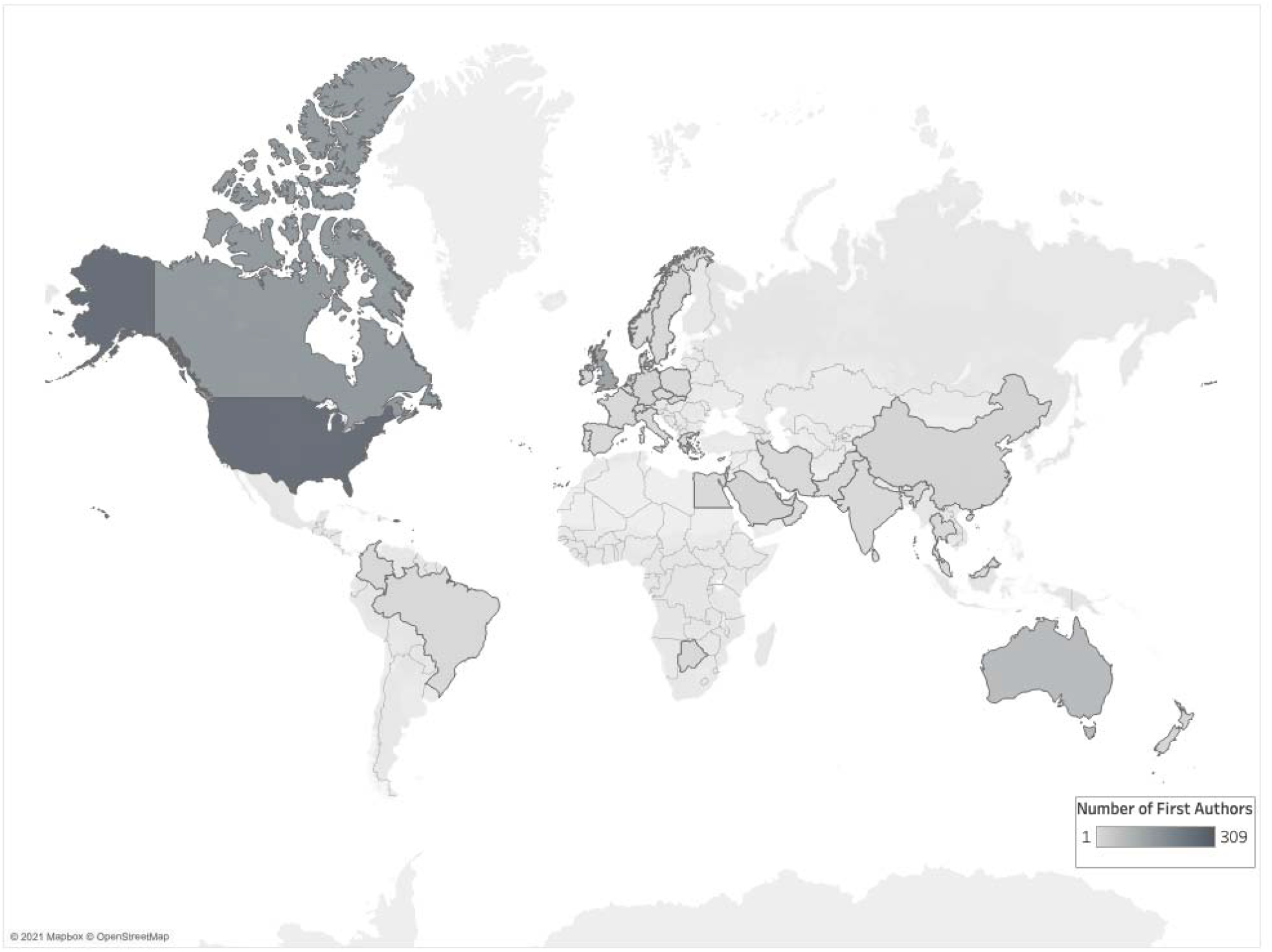
Map highlighting the 42 countries listed as affiliations of first authors of knowledge syntheses published in 14 core medical education journals published between 1999-2019

### Institutions

Across all authorship positions, we identified 617 unique institutions (See Supplemental Appendix D for complete list of institutions). Institutions most often represented were the University of Toronto (n=212 authors), the Mayo Clinic (n=110 authors), and Maastricht University (n=89 authors). See Table 1 for the top 10 institutions by frequency. Nearly half (n=451; 47%) of knowledge synthesis listed authors from a single institution. The most institutions represented on a knowledge synthesis was 14.^34^

**Table 1:** Top 10 institutional affiliations by count of first authors of knowledge syntheses published in 14 core medical education journals published between 1999-2019.

| Institution (country) | Count of first authors (%) | Times Higher Education Ranking | Count of authors across all authorship positions (%) |
| --- | --- | --- | --- |
| University of Toronto (Canada) | 56 (5.7) | 18 | 212 (5.2) |
| Mayo Clinic (United States) | 32 (3.3) | Unranked | 110 (2.7) |
| McMaster University (Canada) | 18 (1.9) | 72 | 64 (1.6) |
| Monash University (Australia) | 18 (1.9) | 75* | 72 (1.8) |
| University of Ottawa (Canada) | 18 (1.9) | 141* | 72 (1.8) |
| National Health Service (United Kingdom) | 17 (1.8) | Unranked | 70 (1.7) |
| University of British Columbia (Canada) | 15 (1.6) | 34 | 52 (1.3) |
| University of Utrecht (Netherlands) | 14 (1.5) | 75* | 76 (1.8) |
| McGill University (Canada) | 13 (1.3) | 42 | 52 (1.3) |
| University of Alberta (Canada) | 13 (1.3) | 136* | 54 (1.3) |
| University of Calgary (Canada) | 13 (1.3) | 201- 250 | 47 (1.1) |
| Maastricht University (Netherlands) | 13 (1.3) | 127 | 89 (2.2) |

Rankings retrieved from *Times Higher Education* (THE) World University Rankings 2020.
\* indicates a tie in THE rankings

For all authorship positions, 78% (n=753) of knowledge syntheses included authors from institutions ranked in the THE top 200. The remaining knowledge syntheses included 368 with authors based at institutions ranked between 200-1000 and 217 at institutions ranked below 1000 or unranked. Two hundred and twenty-nine authors (6%) represented non-university affiliated medical centers or hospitals and 229 (6%) listed affiliations at professional organizations.

First authors represented 362 unique institutions with the most first authors based at the University of Toronto (n=56, 15%) and the Mayo Clinic (n=31, 9%). The third most frequently represented institution was a tie between Monash University, McMaster University, University of Ottawa, and the National Health Service (n=18; 5%). Of the top 200 institutions, only 110 (30%) institutions were represented in the first author position, yet the top 200 institutions accounted for 486 (50%) publications in our sample. Beyond the top 200, first authors represented 101 (28%) institutions ranked between 200-1000 and another 89 (25%) institutions were beyond 1000 or unranked. Thirty-four (9%) first authors represented professional organizations and 25 (7%) non-university affiliated medical centers or hospitals.

## Discussion

The majority of knowledge syntheses were multi-authored manuscripts, which aligns with the description of medical education researchers as inherently collaborative^35^ and may reflect the significant effort required to publish a rigorous knowledge synthesis.^36^ Notably, over the 20 years examined here, the average size of the author team increased. This growth mirrors a similar trend in science more broadly^37^ and in some medical specialties more specifically.^38,39^ Researchers have attributed this growth to multiple factors, including increased ease of collaboration between scholars as a result of computer and internet technology; utilization of research methodologies that require a variety of expertise and skills; and the growing complexity of topics/research questions addressed.^40,41^ In addition, findings from a study of research practices in medical education suggest that some of this growth in author team size may be related to questionable research practices like honorary authorship.^42,43^ As the field of medical education continues to mature and its literature base grows, strategic selection of team members, including the rightsizing of the knowledge synthesis team, will become increasingly important. Thus, future research may be needed to further examine the composition and size of knowledge synthesis teams in order to provide evidence-based practical guidance on team construction.

The majority of author teams included both genders, suggesting that both male and female voices are present across the evidence base being synthesized by these reviews. Over the study period, the ratio of females to males has increased. Additionally, gender parity among authorship teams has been achieved in recent years, which aligns with Madden’s recent analysis of four medical education journals across a variety of publication types.^21^ Furthermore, similar to the findings from Madden et al examining other medical specialities,^44-46^ we identified significantly more males in the last author position. In biomedicine, the last position is traditionally occupied by the senior author who takes on a leadership role in the study^47^ or is often the principal investigator of the research laboratory conducting the work. Similarly, in medical education, the last author position is often held by the “senior author.” While further investigations are necessary, we propose that this finding may be related to the under representation of women in leadership positions in academic medicine.^48^ Overall, however, these gender results are encouraging; nonetheless, future work should continue to track author gender in medical education to monitor for additional changes that might occur over time. For example, recent research related to the impact of the COVID-19 pandemic on science raises concern that female investigators, especially those with younger children, have had less time for research^49^ and writing and, as a result, may be publishing fewer papers than their male counterparts.^50^

Although we identified author representation from 58 countries, 80% of authors teams were based in a single country. Additionally, authorship was dominated by individuals based in the US, Canada, and the UK, suggesting a heavy influence from English-speaking countries. This may have implications for the inclusion or potential exclusion of non-English language articles from reviews, as multinational teams are more likely to include non-English studies in their knowledge syntheses.^49^ The exclusion of non-English articles is a known issue in the conduct of knowledge syntheses and has been labeled the “Tower of Babel Bias”;^52^ this bias has implications for the accuracy and generalizability of research findings.^53^ While examining the language inclusion criteria of each article is beyond the scope of this study, our findings suggest further investigation is warranted to better understand if the Tower of Babel Bias is an important issue in medical education knowledge syntheses.

Few authors listed affiliations in LMIC, and there was even less representation from LMIC in the first authorship position. This suggests geographical diversity is lacking in medical education knowledge syntheses, which has implications for the relevance of these reviews. It also raises questions about potential research waste and bias.^19^ In a 2019 study with similar findings to the present investigation, Thomas concluded that medical education research, more than any other field, is conducted by authors in the English-speaking Western countries, which he referred to as the “realm of the rich”.^54^ This dominance of authors based in Western countries may limit the utility of these findings for non-Western health and education systems. To address some of this imbalance, the Cochrane Collaboration suggests that knowledge syntheses authors “take account of the needs of resource-poor countries and regions in the review process and invite appropriate input on the scope of the review and the questions it will address.”^55^

Researchers have identified that knowledge syntheses conducted across multiple institutions can improve the quality and visibility of a publication, as well as help to avoid a “silo effect”.^56,57^ Our findings demonstrate that just under half of the knowledge syntheses examined were multi-institutional investigations. Moreover, across all authorship positions, 78% of knowledge syntheses included authors affiliated with institutions ranked in the THE Top 200. As other medical education researchers have noted “The big players are in a position to influence the global discourse more than others”.^58^ As such, the field would do well to consider growing the number of multi-institutional collaborations that not only perform empirical research, but also collaborate to conduct knowledge syntheses.

### Limitations and Future Directions

There are a number of important limitations in the present work that suggest some fruitful areas of future research. First, our data set is composed of 14 core journals, which did not include journals from specific world regions, such as the *African Journal of Health Professions Education*. This limitation is particularly important, especially because we wanted to understand who does and does not have a voice in the development of knowledge syntheses. Had the data set examined other journals, we would likely have attained different results. That said, these 14 journals have been defined earlier as core medical education publications.^59,60^

Second, we used a gender prediction tool to determine whether a first name was characterized as male or female. We recognize that this is an oversimplification of a complicated social construct like gender, especially because an individual’s gender is best described by that individual. However, this is a starting point necessary to begin to provide a sense of the field and follows the protocol of similar papers published on this topic.^21,61^

Third, we did not review the full-texts of the knowledge syntheses we analyzed. Therefore, we are unable to make any claims on how author characteristics may have impacted conduct of the knowledge synthesis or their conclusions. Future work should consider a more in-depth examination of the full text to examine components like inclusion and exclusion criteria and the stated rationale behind the authors’ decisions. This future work might also include qualitative inquiry to better understand how authors approached their review, including reflections on how their backgrounds may have impacted the conduct of the review. However, we did identify knowledge syntheses from 58 countries, including representation from all World Bank regions. In relation to THE ranking, not all universities submit data for ranking and thus institutions may have been missed. Additionally, we identified authors from associations and organizations, which would not have been ranked, but that may have influence (e.g., the Association of American Medical Colleges).

Finally, our research team is made up of individuals from the US, Canada, and the Netherlands. The irony of this team composition is not lost on us, and we certainly acknowledge our privileged positions, which likely limit our ability to appreciate the various world perspectives that may be relevant in understanding important author characteristics.

### Conclusion

The production of knowledge syntheses, like all knowledge production, can be influenced by the authors’ characteristics, backgrounds, and the power structures and cultural norms from which they operate.^9,19^ In this study, we identified and critically examined the characteristics of the authors of knowledge syntheses to better understand the voices present – and those that may be missing – in our evidence base. While gender parity has improved in recent years, knowledge synthesis authors predominantly work in elite institutions from high-income countries. Although more research is needed to truly understand the impact of these and other author characteristics, we suspect that some of the imbalances observed herein may have negative implications for medical education’s evidence base and its global relevance.

## Data Availability

This study utilized the following publicly available data set: Maggio LA, Costello J, Norton C, Driessen EW, Artino AR. Knowledge synthesis in medical education: A bibliometric analysis [data set]. 2020. Zenodo: http://doi.org/10.5281/zenodo.3990481.

## References

1. Harden RM, Grant J, Buckley G, Hart IR. Best evidence medical education. Adv Health Sci Educ Theory Pract. 2000;5(1):71–90.

2. Gordon M. Are we talking the same paradigm? Considering methodological choices in health education systematic review. Med Teach. 2016;38(7):746–750.

3. Maggio LA, Costello JA, Norton C, Driessen EW, Artino AR Jr. Knowledge syntheses in medical education: A bibliometric analysis [published online ahead of print, Oct 22, 2020]. Perspect Med Educ. 2020;1–9. DOI:10.1007/s40037-020-00626-9.

4. Harden RM, Grant J, Buckley G, Hart IR. BEME Guide No. 1: Best Evidence Medical Education. Med Teach. 1999;21(6):553–562.

5. Tricco AC, Tetzlaff J, Moher D. The art and science of knowledge synthesis. Journal of clinical epidemiology. 2011;64(1):11–20.

6. Cook D, West C. Conducting systematic reviews in medical education: a stepwise approach. Med Educ. 2012;46(10):943–52.

7. Maggio LA, Thomas A, Durning SJ. Knowledge Synthesis. In: Understanding Medical Education: Evidence, Theory, and Practice. Chichester, UK: John Wiley & Sons; 2018; 457–469.

8. Best Evidence Medical Education. Forming a review group. Available from: https://www.bemecollaboration.org/Step+2+Review+Group/. Accessed January, 2021.

9. Snyder H. Literature review as a research methodology: an overview and guidelines. Journal of Business Research. 2019;104:333–339.

10. Holman L, Stuart-Fox D, Hauser CE. The gender gap in science: how long until women are equally represented? PLoS biology. 2018;16(4):e2004956.

11. Macaluso B, Larivière V, Sugimoto T, Sugimoto CR. Is science built on the shoulders of women? A study of gender differences in contributorship. Acad Med. 2016;91(8):1136–1142.

12. Raj A, Carr PL, Kaplan SE, Terrin N, Breeze JL, Freund KM. Longitudinal analysis of gender differences in academic productivity among medical faculty across 24 medical schools in the United States. Acad Med. 2016;91(8):1074–1079.

13. Lundine J, Bourgeault IL, Clark J, Heidari S, Balabanova D. The gendered system of academic publishing. Lancet. 2018;391(10132):1754–1756.

14. Kelaher M, Ng L, Knight K, Rahadi A. Equity in global health research in the new millennium: trends in first-authorship for randomized controlled trials among low-and middle-income country researchers 1990-2013. Int J Epidemiol. 2016;45(6):2174–83.

15. Bhandal T. Ethical globalization? Decolonizing theoretical perspectives for internationalization in Canadian medical education. Can Med Educ J. 2018;9(2):e33–45

16. Harris M, Marti J, Watt H, Bhatti Y, Macinko J, Darzi A. Explicit bias toward high-income-country research: a randomized, blinded, crossover experiment of English clinicians. Health Aff. 2017;36(11):1997–2004.

17. Walker R, Barros B, Conejo R, Neumann K, Telefont M. Personal attributes of authors and reviewers, social bias and the outcomes of peer review: a case study. F1000Res. 2015;4:21.

18. Skopec M, Issa H, Reed J, Harris M. The role of geographic bias in knowledge diffusion: a systematic review and narrative synthesis. Res Integr Peer Rev. 2020;5:2.

19. Higgins JPT, Thomas J, Chandler J, Cumpston M, Li T, Page MJ, Welch VA (editors). Cochrane Handbook for Systematic Reviews of Interventions version 6.1 (updated September 2020). Cochrane, 2020. Available from www.training.cochrane.org/handbook.

20. Cochrane. Cochrane Strategy, 2020. https://www.cochrane.org/about-us/strategy-to-2020. accessed January, 2021.

21. Madden C, O’Malley R, O’Connor P, O’Dowd E, Byrne D, Lydon S. Gender in authorship and editorship in medical education journals: A bibliometric review [published online ahead of print, Dec 1, 2020]. Med Educ. DOI:10.1111/medu.14427.

22. Thomas MP. The geographic and topical landscape of medical education research. BMC Med Educ. 2019;19(1):189.

23. Maggio LA, Costello J, Norton C, Driessen EW, Artino AR. Knowledge synthesis in medical education: A bibliometric analysis [data set]. 2020. Zenodo: http://doi.org/10.5281/zenodo.3990481.

24. Federer LM, Lu YL, Joubert DJ, Welsh J, Brandys B. Biomedical Data Sharing and Reuse: Attitudes and Practices of Clinical and Scientific Research Staff. PLoS One. 2015;10(6):e0129506.

25. Perrier L, Blondal E, MacDonald H. The views, perspectives, and experiences of academic researchers with data sharing and reuse: A meta-synthesis [published correction appears in PLoS One. 2020;15(6):e0234275.. PLoS One. 2020;15(2):e0229182.

26. Google Sheets. Google LLC. 2020.

27. Demografix ApS. genderize.io. https://genderize.io/. accessed December, 2020.

28. Hart KL, Perlis RH. Trends in proportion of women as authors of medical Journal articles, 2008-2018. JAMA Intern Med. 2019;179(9):1285–1287.

29. Bagga E, Stewart S, Gamble GD, Hill J, Grey A, Dalbeth N. Representation of Women as Authors of Rheumatology Research Articles [published online ahead of print, Aug 18, 2020]. Arthritis Rheumatol. DOI:10.1002/art.41490.

30. The World Bank. World Bank Country and Lending Groups. http://databank.worldbank.org/data/download/site-content/CLASS.xls. accessed February, 2021.

31. Times Higher Education. Times Higher Education World University Rankings 2020. https://www.timeshighereducation.com/world-university-rankings/2020/world-ranking#!/page/0/length/-1/sort_by/rank/sort_order/asc/cols/scores. accessed December, 2020.

32. Tableau. Tableau Public. Seattle (WA):Tableau;c2020. Available from: https://public.tableau.com/en-us/s/download.

33. Milestone Consortium, Russet F, Humbertclaude V, et al. Training of adult psychiatrists and child and adolescent psychiatrists in europe: a systematic review of training characteristics and transition from child/adolescent to adult mental health services. BMC Med Educ. 2019;19(1):204.

34. Daniel M, Rencic J, Durning SJ, et al. Clinical Reasoning Assessment Methods: A Scoping Review and Practical Guidance. Acad Med. 2019;94(6):902–912.

35. O’Sullivan P, Stoddard H, Kalishman S. Collaborative research in medical education: a discussion of theory and practice. Med Educ. 2010;44(12):1175–1184

36. Khangura S, Konnyu K, Cushman R, Grimshaw J, Moher D. Evidence summaries: the evolution of a rapid review approach. Syst Rev. 2012;1:10.

37. Baethge C. Publish together or perish: the increasing number of authors per article in academic journals is the consequence of a changing scientific culture. Some researchers define authorship quite loosely. Dtsch Arztebl Int. 2008;105(20):380–383.

38. Dang W, McInnes MD, Kielar AZ, Hong J. A Comprehensive Analysis of Authorship in Radiology Journals [published correction appears in PLoS One. 2016;11(1):e0147166.. PLoS One. 2015;10(9):e0139005.

39. Lammers R, Simunich T, Ashurst J. Authorship trends of emergency medicine publications over the last two decades. West J Emerg Med. 2016;17(3):367–371.

40. Fontanarosa P, Bauchner H, Flanagin A. Authorship and Team Science. JAMA. 2017;318(24):2433–2437.

41. Tilak G, Prasad V, Jena AB. Authorship Inflation in Medical Publications. Inquiry. 2015;52:0046958015598311.

42. Artino AR Jr, Driessen EW, Maggio LA. Ethical Shades of Gray: International Frequency of Scientific Misconduct and Questionable Research Practices in Health Professions Education. Acad Med. 2019;94(1):76–84.

43. Maggio LA, Artino AR Jr, Watling CJ, Driessen EW, O’Brien BC. Exploring researchers’ perspectives on authorship decision making. Med Educ. 2019;53(12):1253–1262.

44. Webb J, Cambron J, Xu KT, Simmons M, Richman P. First and last authorship by gender in emergency medicine publications-a comparison of 2008 vs. 2018 [published online ahead of print, Oct 28, 2020]. Am J Emerg Med. 2020. DOI:10.1016/j.ajem.2020.10.045

45. Fishman M, Williams II WA, Goodman DM, Ross LF. Gender differences in the authorship of original research in pediatric journals, 2001-2016. J Pediatr. 2017;191:244-249.e1

46. Vranas KC, Ouyang D, Lin AL, et al. Gender differences in authorship of critical care literature. Ame J Respir Crit Care Med. 2020;201(7):840–847.

47. Smith E, Williams-Jones B. Authorship and responsibility in health sciences research: a review of procedures for fairly allocating authorship in multi-author studies. Sci Eng Ethics. 2012;18(2):199–212.

48. Carr PL, Gunn CM, Kaplan SA, Raj A, Freund KM. Inadequate progress for women in academic medicine: findings from the National Faculty Study. J Womens Health (Larchmt). 2015;24(3):190–199.

49. Deryugina T, Shurchkov O, Stearns JE. Covid-19 disruptions disproportionately affect female academics. https://www.nber.org/papers/w28360 National Bureau of Economic Research working paper 28360. Published January, 2021. accessed January, 2021.

50. Viglione, G. Are women publishing less during the pandemic? Here’s what the data says. Nature. 2020;581;365–366.

51. Neimann Rasmussen L, Montgomery P. The prevalence of and factors associated with inclusion of non-English language studies in Campbell systematic reviews: a survey and meta-epidemiological study. Syst Rev. 2018;7(1):129.

52. Gregoire G, Derderian F, Le LJ. Selecting the language of the publications included in a meta-analysis: is there a Tower of Babel bias? J Clin Epidemiol 1995;48(1):159–163.

53. Jackson JL, Kuriyama A. How Often Do Systematic Reviews Exclude Articles Not Published in English? J Gen Intern Med. 2019;34(8):1388–1389.

54. Thomas MP. The geographic and topical landscape of medical education research. BMC Med Educ. 2019;19(1):189.

55. Lasserson TJ, Thomas J, Higgins JPT. Chapter 1: Starting a review. In: Higgins Jpt, Thomas J, Chandler J, Cumpston M, Li T, Page MJ, Welch VA (editors). Cochrane Handbook for Systematic Reviews of Interventions version 6.1 (updated September 2020). Cochrane, 2020. Available from www.training.cochrane.org/handbook.

56. Catalá-López F, Alonso-Arroyo A, Hutton B, Aleixandre-Benavent R, Moher D. Global collaborative networks on meta-analyses of randomized trials published in high impact factor medical journals: a social network analysis. BMC Med. 2014;12:15.

57. Uttley L, Montgomery P. The influence of the team in conducting a systematic review. Syst Rev. 2017;6(1):149.

58. Frambach JM, Martimianakis MA. The discomfort of an educator’s critical conscience: the case of problem-based learning and other global industries in medical education. Perspect Med Educ. 2017;6(1):1–4.

59. Maggio LA, Meyer HS, Artino AR Jr.. Beyond citation rates: a real-time impact analysis of health professions education research using altmetrics. Acad Med. 2017;92(10):1449–1455.

60. Lee K, Whelan JS, Tannery NH, Kanter SL, Peters AS. 50 years of publication in the field of medical education. Med Teach. 2013;35:591–8.

61. Mamtani M, Shofer F, Mudan A, et al. Quantifying gender disparity in physician authorship among commentary articles in three high-impact medical journals: an observational study. BMJ Open. 2020;10(2):e034056.

