## Supplemental Appendix A for "Knowledge syntheses in medical education: Examining author gender, geographic location, and institutional affiliation"

Core medical education journals (n=14) included

*Academic Medicine, Advances in Health Sciences Education, BMC Medical Education, Canadian Medical Education Journal, Clinical Teacher, International Journal of Medical Education, Advances in Medical Education and Practice, Journal of Graduate Medical Education, Medical Education, Medical Education Online, Medical Teacher, Perspectives on Medical Education, Teaching and Learning in Medicine*, and *The Journal of Continuing Education in the Health Professions*.
