## Supplemental Appendix B for "Knowledge syntheses in medical education: Examining author gender, geographic location, and institutional affiliation"

Supplemental Appendix B: Top 10 most prolific authors overall, first authors, and last authors in publishing knowledge syntheses (KS) in 14 core medical education journals between 1999-2019.

| Most prolific overall authors (*gender) | Number KS | Most prolific first authors *(gender) | Number KS | Most prolific last authors (*gender) | Number KS |
| --- | --- | --- | --- | --- | --- |
| Cook, D (M) | 36 | Cook, D (M) | 21 | Cook, D (M) | 9 |
| Ten Cate, O (M) | 15 | Mcgaghie, W (M) | 6 | Oswald, A (F) | 6 |
| Hatala, R (F) | 12 | Gordon, M (M) | 5 | Ten Cate, O (M) | 6 |
| Scherpbier, A (M) | 12 | Norman, G (M) | 5 | Dornan, T (M) | 4 |
| Mcgaghie, W (M) | 11 | Maudsley, G (F) | 4 | Durning, S (M) | 4 |
| Brydges, R. (M) | 11 | Hauer, K (F) | 4 | Hatala, R (F) | 4 |
| Van Der Vleuten, C (M) | 11 | Burgess, A (F) | 4 | Krishna, L (M) | 4 |
| Dornan, T (M) | 11 | Brydges, R (M) | 4 | Van Der Vleuten, C (M) | 4 |
| Eva, K (M) | 10 | Buckley, S (F) | 3 | Archer, J (M) | 3 |
| Thistlethwaite, J (F) | 9 | Benbassat, J (M) | 3 | Bordage, G (M) | 3 |
|  |  |  |  | Charlin, B (M) | 3 |
|  |  |  |  | Fernandez, R (F) | 3 |
|  |  |  |  | Horsley, T (F) | 3 |
|  |  |  |  | Hu, W (F) | 3 |
|  |  |  |  | Khan, K (M) | 3 |
|  |  |  |  | Lingard, L (F) | 3 |
|  |  |  |  | Maloney, S (M) | 3 |
|  |  |  |  | McKelvy, D (F) | 3 |
|  |  |  |  | Mellis, C (M) | 3 |
|  |  |  |  | O'Sullivan, P (F) | 3 |
|  |  |  |  | Reeves, S (M) | 3 |
|  |  |  |  | Roberts, T (F) | 3 |
|  |  |  |  | Scalese, R (M) | 3 |
|  |  |  |  | Scherpbier, A (M) | 3 |
|  |  |  |  | Schuwirth, L (M) | 3 |
|  |  |  |  | Thistlethwaite, J (F) | 3 |
|  |  |  |  | van Merrienboer, J (M) | 3 |
|  |  |  |  | Violato, C (M) | 3 |
|  |  |  |  | Wayne, D (F) | 3 |
|  |  |  |  | Wieringa-de Waard, M (F) | 3 |
|  |  |  |  | Williams,Brett (M) | 3 |

*Gender: M=male, F=female
