## Supplemental Appendix C for "Knowledge syntheses in medical education: Examining author gender, geographic location, and institutional affiliation"

Appendix C: Country affiliations for all and first authors of knowledge syntheses (KS) published in 14 core medical education journals between 1999-2019.

| Country | Count of KS with representation in any author position | Count of KS with representation by first author | World Bank Country Classification |
| --- | --- | --- | --- |
| Argentina | 1 | 0 | Low & middle income |
| Australia | 111 | 85 | High income |
| Austria | 1 | 0 | High income |
| Bahrain | 5 | 4 | High income |
| Barbados | 1 | 0 | High income |
| Belgium | 13 | 8 | High income |
| Botswana | 1 | 1 | Low & middle income |
| Brazil | 4 | 3 | Low & middle income |
| Canada | 233 | 183 | High income |
| Chile | 2 | 0 | High income |
| China | 22 | 19 | Low & middle income |
| Colombia | 1 | 1 | Low & middle income |
| Croatia | 1 | 0 | High income |
| Cyprus | 1 | 1 | High income |
| Czech Republic | 1 | 1 | High income |
| Denmark | 8 | 8 | High income |
| Dominica | 1 | 1 | Low & middle income |
| Egypt | 3 | 1 | Low & middle income |
| France | 4 | 4 | High income |
| Germany | 19 | 16 | High income |
| Ghana | 1 | 0 | Low & middle income |
| Greece | 2 | 2 | High income |
| Grenada | 1 | 1 | Low & middle income |
| Hungary | 1 | 0 | High income |
| India | 4 | 2 | Low & middle income |
| Iran | 4 | 4 | Low & middle income |
| Ireland | 14 | 10 | High income |
| Israel | 4 | 3 | High income |
| Italy | 5 | 1 | High income |
| Jordan | 1 | 0 | Low & middle income |
| Kuwait | 1 | 0 | High income |
| Lithuania | 1 | 0 | High income |
| Malaysia | 1 | 1 | Low & middle income |
| Nigeria | 1 | 0 | Low & middle income |
| Netherlands | 105 | 65 | High income |
| Norway | 4 | 3 | High income |
| New Zealand | 16 | 12 | High income |
| Oman | 1 | 1 | High income |
| Pakistan | 2 | 1 | Low & middle income |
| Philippines | 1 | 0 | Low & middle income |
| Poland | 3 | 1 | High income |
| Portugal | 5 | 4 | High income |
| Qatar | 2 | 1 | High income |
| Saudi Arabia | 20 | 18 | High income |
| Singapore | 17 | 15 | High income |
| Slovakia | 6 | 5 | High income |
| Spain | 3 | 1 | High income |
| Sri lanka | 1 | 1 | Low & middle income |
| Sweden | 4 | 3 | High income |
| Switzerland | 10 | 5 | High income |
| Tanzania | 1 | 0 | Low & middle income |
| Thailand | 4 | 3 | Low & middle income |
| Turkey | 1 | 0 | Low & middle income |
| United Arab Emirates | 3 | 1 | High income |
| Uganda | 1 | 0 | Low & middle income |
| United Kingdom | 180 | 151 | High income |
| United States | 366 | 312 | High income |
| Vietnam | 1 | 0 | Low & middle income |
