## Supplemental Appendix D for "Knowledge syntheses in medical education: Examining author gender, geographic location, and institutional affiliation"

Appendix D: Institutional affiliation and Times Higher Education (THE) ranking of knowledge syntheses (KS) published in 14 core medical education journals between 1999-2019.

| THE rank or classification | Institution | Count of KS across all author positions |
| --- | --- | --- |
| 1 | Univ Oxford | 17 |
| 3 | Univ Cambridge | 21 |
| 4 | Stanford | 16 |
| 7 | Harvard | 30 |
| 8 | Yale | 16 |
| 9 | Univ Chicago | 14 |
| 10 | Imperial Coll | 12 |
| 11 | Univ Penn | 8 |
| 12 | Johns Hopkins Univ | 67 |
| 15 | Univ College London | 35 |
| 16 | Columbia Univ | 6 |
| 17 | Univ California Los Angeles | 22 |
| 18 | Univ Toronto | 212 |
| 19 | Cornell Univ | 16 |
| 20 | Duke Univ | 38 |
| 21 | Univ Michigan | 29 |
| 22 | Northwestern Univ | 34 |
| 24 | Peking Univ | 2 |
| 25 | Natl Univ Singapore | 35 |
| 26 | Univ Washington | 44 |
| 29 | New York Univ | 21 |
| 30 | Univ Edinburgh | 7 |
| 32 | Univ Melbourne | 9 |
| 32 | Univ Munich | 15 |
| 34 | Univ British Columbia | 52 |
| 35 | Univ Hong Kong | 3 |
| 36 | Kings Coll | 17 |
| 38 | Univ Texas Austin | 1 |
| 42 | McGill Univ | 52 |
| 45 | Univ Leuven | 6 |
| 48 | Nanyang Tech | 4 |
| 50 | Australian Natl Univ | 1 |
| 51 | Univ Wisconsin | 17 |
| 52 | Washington Univ | 5 |
| 53 | Brown Univ | 9 |
| 54 | Univ No Carolina | 13 |
| 55 | Univ California Davis | 7 |
| 55 | Univ Manchester | 11 |
| 60 | Univ Sydney | 27 |
| 61 | Boston Univ | 7 |
| 62 | Univ Amsterdam | 35 |
| 62 | Univ Southern Calif | 8 |
| 66 | Univ Queensland | 30 |
| 67 | Delft University | 2 |
| 67 | Leiden Univ | 2 |
| 69 | Erasmus Univ | 15 |
| 70 | Ohio State Univ | 10 |
| 72 | McMaster Univ | 64 |
| 73 | Univ Groningen | 16 |
| 75 | Monash Univ | 72 |
| 75 | Royal Dutch Med | 3 |
| 75 | Univ Utrecht | 76 |
| 77 | Univ Warwick | 23 |
| 77 | Warwick | 9 |
| 78 | Penn State | 13 |
| 79 | Univ Minnesota | 6 |
| 80 | Charite | 4 |
| 80 | Emory Univ | 3 |
| 84 | Mich St Univ | 10 |
| 85 | Univ Montreal | 17 |
| 87 | Queens Univ Belfast | 5 |
| 87 | Univ Bristol | 4 |
| 90 | Univ Zurich | 5 |
| 91 | Univ Tübingen | 17 |
| 94 | Dartmouth | 4 |
| 94 | Univ Basel | 1 |
| 96 | Univ Calif - Irvine | 2 |
| 99 | Univ Glasgow | 2 |
| 101 | Univ Copenhagen | 8 |
| 102 | Georgetown Univ | 3 |
| 104 | Univ Arizona | 7 |
| 107 | Univ Virginia | 11 |
| 110 | Queen Mary Univ | 4 |
| 112 | Univ Birmingham | 34 |
| 113 | Univ Bern | 6 |
| 113 | Univ Pittsburgh | 17 |
| 115 | Aarhus | 8 |
| 116 | Vanderbilt Univ | 10 |
| 117 | Univ Sheffield | 9 |
| 119 | Case Western | 3 |
| 120 | Natl Taiwan | 7 |
| 120 | Univ Adelaide | 2 |
| 122 | Univ Southampton | 2 |
| 123 | Ghent Univ | 12 |
| 124 | Univ Colorado | 1 |
| 125 | Univ Göttingen | 2 |
| 127 | Maastricht Univ | 89 |
| 128 | Radboud | 24 |
| 130 | Univ Paris | 1 |
| 131 | Univ Oslo | 1 |
| 131 | Univ W Australia | 1 |
| 133 | Univ Durham | 8 |
| 134 | Indiana Univ | 14 |
| 136 | Univ Alberta | 54 |
| 136 | Univ Cape Town | 2 |
| 138 | Vrije Univ | 31 |
| 139 | Tufts Univ | 26 |
| 139 | Univ Lancaster | 1 |
| 141 | Ulm Univ | 1 |
| 141 | Univ Ottawa | 72 |
| 144 | Nanjing Med | 17 |
| 144 | Univ Geneva | 2 |
| 146 | Univ Exeter | 7 |
| 149 | Univ Hamburg | 3 |
| 152 | Catholic Univ Louvain | 6 |
| 152 | Univ Nottingham | 11 |
| 155 | Univ Leeds | 24 |
| 157 | Shanghai Jiao Tong | 8 |
| 157 | TU Dresden | 2 |
| 157 | Univ Auto Barcelona | 1 |
| 157 | Univ Cologne | 1 |
| 164 | Trinity Coll Dublin | 5 |
| 165 | Univ Liverpool | 25 |
| 166 | Leicester University | 2 |
| 166 | Univ Leicester | 13 |
| 168 | Rutgers | 6 |
| 168 | Univ Aberdeen | 8 |
| 172 | Univ Alabama | 16 |
| 173 | Univ Rochester | 5 |
| 178 | Texas A&M Univ | 1 |
| 179 | Auckland Univ | 1 |
| 179 | Korea Univ | 1 |
| 179 | Queensland Univ Technol | 3 |
| 179 | Univ Auckland | 23 |
| 186 | Eindhoven Univ Technol | 1 |
| 192 | Univ E Anglia | 10 |
| 194 | Univ Technol Sydney | 6 |
| 197 | Yonsei Univ | 3 |
| 198 | Cardiff Univ | 12 |
| 198 | George Washington Univ | 10 |
| 198 | Univ Antwerp | 9 |
| 198 | Univ Cardiff | 1 |
| 198 | Univ Lausanne | 1 |
| 201 - 250 | Griffith | 14 |
| 201 - 250 | Howard Univ | 3 |
| 201 - 250 | James Cook Univ | 8 |
| 201 - 250 | King Abdulaziz | 3 |
| 201 - 250 | King Saud Bin | 7 |
| 201 - 250 | Macquarie | 7 |
| 201 - 250 | Med Univ Graz | 1 |
| 201 - 250 | Newcastle Univ | 3 |
| 201 - 250 | Qatar Univ | 1 |
| 201 - 250 | Univ Calgary | 47 |
| 201 - 250 | Univ Coll Dublin | 3 |
| 201 - 250 | Univ Dundee | 11 |
| 201 - 250 | Univ Iowa | 5 |
| 201 - 250 | Univ Massachusetts | 16 |
| 201 - 250 | Univ Miami | 14 |
| 201 - 250 | Univ Otago | 23 |
| 201 - 250 | Univ So Florida | 2 |
| 201 - 250 | Univ Twente | 1 |
| 201 - 250 | Univ Utah | 3 |
| 201 - 250 | Univ Waterloo | 2 |
| 201 - 250 | Univ Wollongon | 8 |
| 201 - 250 | Wake Forest | 20 |
| 201 - 250 | Western Univ | 31 |
| 251 - 300 | Brighton & Sussex | 8 |
| 251 - 300 | Curtin Univ | 1 |
| 251 - 300 | Dalhousie | 12 |
| 251 - 300 | Flinders | 36 |
| 251 - 300 | Florida St | 1 |
| 251 - 300 | La Trobe Univ | 2 |
| 251 - 300 | Natl Univ Ireland | 16 |
| 251 - 300 | Ore Hlth St Univ | 11 |
| 251 - 300 | Queen's Univ Kingston | 20 |
| 251 - 300 | Stellenbosch | 16 |
| 251 - 300 | Swansea Univ | 2 |
| 251 - 300 | Univ Auto Madrid | 1 |
| 251 - 300 | Univ Cincinnati | 1 |
| 251 - 300 | Univ Essex | 1 |
| 251 - 300 | Univ Illinois | 35 |
| 251 - 300 | Univ Laval | 9 |
| 251 - 300 | Univ Sao Paulo | 1 |
| 251 - 300 | Univ So Australia | 17 |
| 251 - 300 | Univ So Denmark | 5 |
| 251 - 300 | Univ W Sydney | 9 |
| 251 - 300 | Western Sydney Univ | 9 |
| 252 - 300 | Univ Illinois Chi | 7 |
| 301 - 350 | Deakin Univ | 8 |
| 301 - 350 | Goethe Univ | 3 |
| 301 - 350 | NC St Univ | 2 |
| 301 - 350 | Rush Univ | 3 |
| 301 - 350 | SUNY Stony Brook | 2 |
| 301 - 350 | Temple Univ | 4 |
| 301 - 350 | Tulane | 2 |
| 301 - 350 | Univ Coll Cork | 3 |
| 301 - 350 | Univ Colo Denver | 1 |
| 301 - 350 | Univ Fribourg | 1 |
| 301 - 350 | Univ Montpellier | 2 |
| 301 - 350 | Univ New Mexico | 4 |
| 301 - 350 | Univ Newcastle | 16 |
| 301 - 350 | Univ Tasmania | 5 |
| 301 - 350 | Univ Tennessee | 6 |
| 351 - 400 | Aust Cath Univ | 2 |
| 351 - 400 | Northumbria Univ | 2 |
| 351 - 400 | SUNY Albany | 1 |
| 351 - 400 | Swinburne Univ | 1 |
| 351 - 400 | Univ Connecticut | 2 |
| 351 - 400 | Univ Crete | 3 |
| 351 - 400 | Univ Manitoba | 2 |
| 351 - 400 | Wayne State Univ | 2 |
| 401 - 500 | Bond Univ | 12 |
| 401 - 500 | Bournemouth | 3 |
| 401 - 500 | City Univ London | 4 |
| 401 - 500 | Drexel | 4 |
| 401 - 500 | Liverpool John Moores | 2 |
| 401 - 500 | Pontificia | 10 |
| 401 - 500 | St Catherines Coll | 1 |
| 401 - 500 | Tongji Univ | 4 |
| 401 - 500 | Univ Bordeaux | 1 |
| 401 - 500 | Univ Georgia | 5 |
| 401 - 500 | Univ Kansas | 2 |
| 401 - 500 | Univ Kentucky | 1 |
| 401 - 500 | Univ KwaZulu | 9 |
| 401 - 500 | Univ Philippines | 1 |
| 401 - 500 | Univ Porto | 2 |
| 401 - 500 | Univ Saskatchewan | 2 |
| 401 - 500 | Univ So Carolina | 6 |
| 401 - 500 | Univ Texas San Antonio | 6 |
| 401 - 500 | Univ Turin | 2 |
| 401 - 500 | York Univ | 6 |
| 501 - 600 | Bar-Ilan Univ | 1 |
| 501 - 600 | Carleton Univ | 1 |
| 501 - 600 | Clau Bern Univ | 1 |
| 501 - 600 | Keele | 10 |
| 501 - 600 | King Saud Univ | 1 |
| 501 - 600 | LSU | 1 |
| 501 - 600 | Mem Univ | 11 |
| 501 - 600 | Natl Yang-Ming Univ | 3 |
| 501 - 600 | Plymouth Univ | 6 |
| 501 - 600 | Suez Canal Univ | 1 |
| 501 - 600 | Tor Vergata Univ | 2 |
| 501 - 600 | Univ Cattolica | 1 |
| 501 - 600 | Univ Complutense | 2 |
| 501 - 600 | Univ Hull | 1 |
| 501 - 600 | Univ Ibadan | 1 |
| 501 - 600 | Univ Keele | 3 |
| 501 - 600 | Univ Limerick | 9 |
| 501 - 600 | Univ Lisbon | 4 |
| 501 - 600 | Univ Nebraska | 6 |
| 501 - 600 | Univ Plymouth | 18 |
| 501 - 600 | Univ Tehran | 1 |
| 501 - 600 | Univ Udine | 2 |
| 501 - 600 | Univ West Indies | 1 |
| 501 - 600 | Xi An Jiao | 7 |
| 601 - 800 | Baylor | 16 |
| 601 - 800 | Coventry Univ | 1 |
| 601 - 800 | Creighton Univ | 5 |
| 601 - 800 | Ewha Womans | 5 |
| 601 - 800 | Fed Univ Sao Paulo | 1 |
| 601 - 800 | Iran Univ | 2 |
| 601 - 800 | Jagiellonian Univ | 1 |
| 601 - 800 | Makerere Univ | 2 |
| 601 - 800 | Masaryk Univ | 2 |
| 601 - 800 | Middle E Tech Univ | 1 |
| 601 - 800 | Old Dominion | 5 |
| 601 - 800 | Pusan Natl Univ | 1 |
| 601 - 800 | Ryerson Univ | 1 |
| 601 - 800 | Shahid Beheshti | 2 |
| 601 - 800 | Sichuan Univ | 19 |
| 601 - 800 | Southern Cross Univ | 5 |
| 601 - 800 | Texas Tech | 5 |
| 601 - 800 | Univ Arkansas | 2 |
| 601 - 800 | Univ Brighton | 1 |
| 601 - 800 | Univ Cent Florida | 5 |
| 601 - 800 | Univ Greenwich | 1 |
| 601 - 800 | Univ Indonesia | 1 |
| 601 - 800 | Univ Lille | 1 |
| 601 - 800 | Univ Maryland | 11 |
| 601 - 800 | Univ Minho | 6 |
| 601 - 800 | Univ Nevada | 2 |
| 601 - 800 | Univ Quebec | 2 |
| 601 - 800 | Univ Sharjah | 1 |
| 601 - 800 | Univ Sherbrooke | 9 |
| 601 - 800 | Univ W Cape | 5 |
| 601 - 800 | Univ Westminster | 1 |
| 601 - 800 | Univ Windsor | 1 |
| 601 - 800 | W Virg Univ | 4 |
| 801 - 1000 | Ain Shams Univ | 1 |
| 801 - 1000 | Birmingham City Univ | 1 |
| 801 - 1000 | Chonnam Natl Univ | 1 |
| 801 - 1000 | Edinburgh Napier | 1 |
| 801 - 1000 | Isfahan Univ | 4 |
| 801 - 1000 | Kingston Univ | 4 |
| 801 - 1000 | Kyungpook Natl Univ | 1 |
| 801 - 1000 | Sultan Qaboos Univ | 1 |
| 801 - 1000 | Univ Bedfordshire | 2 |
| 801 - 1000 | Univ Cent Lancashire | 8 |
| 801 - 1000 | Univ Jordan | 1 |
| 801 - 1000 | Univ No British Columbia | 1 |
| 801 - 1000 | Univ Salford | 2 |
| 801 - 1000 | Univ Szeged | 1 |
| 801 - 1000 | Univ Teesside | 1 |
| 801 - 1000 | Univ Zaragoza | 1 |
| 801 - 1000 | Vilnius Univ | 1 |
| 1001+ | Edge Hill Univ | 4 |
| 1001+ | Hashemite Univ | 1 |
| 1001+ | Oakland Univ | 7 |
| 1001+ | Texas State | 4 |
| 1001+ | UFCSPA | 1 |
| 1001+ | Univ Colombo | 2 |
| 1001+ | Univ Hradec | 2 |
| 1001+ | Univ New South Wales | 9 |
| 1001+ | Univ So Wales | 1 |
|  | Below affiliations were not included in the THE ranking |  |
| IND | Individual | 11 |
| MC | Albert Schweitzer Hosp | 2 |
| MC | Ann Arbor Plast Surg | 1 |
| MC | Army | 8 |
| MC | Assisi Hosp (Singapore) | 2 |
| MC | Atrium | 6 |
| MC | Austin Health | 1 |
| MC | Bahrain Def | 10 |
| MC | Bankstown Campbelltown Hosp | 1 |
| MC | BC Ch & Wom Hosp | 1 |
| MC | Belfast City Hosp | 1 |
| MC | Botsford Gen Hosp | 1 |
| MC | Callen Lorde Comm Hlth Ctr | 1 |
| MC | Cape Fear Valley Hlth Syst | 2 |
| MC | Chang Gung | 7 |
| MC | Christiana | 12 |
| MC | Cleveland Clin | 2 |
| MC | Clin Skills Lab | 1 |
| MC | Cochin Hosp | 2 |
| MC | Emirates Hosp | 1 |
| MC | Evangelismos Med Ctr | 6 |
| MC | Far East Mem Hosp | 1 |
| MC | Hamad Med Corp | 2 |
| MC | HETI | 1 |
| MC | Hop Robert | 1 |
| MC | Hop Univ Jo Rav Andr | 1 |
| MC | Hosp Grp Twente | 1 |
| MC | Hunter New Eng LHD | 1 |
| MC | IAMSPE | 3 |
| MC | Inst Mental Hlth (Singapore) | 11 |
| MC | Jeroen Bosch Hosp | 1 |
| MC | Kaiser Permanente | 2 |
| MC | Kanye 7th Day Adv Hosp | 1 |
| MC | Keesler Med Ctr | 1 |
| MC | KK Womens & Childrens Hosp | 6 |
| MC | Lady Cilento Hosp | 1 |
| MC | Lehigh Valley Hlth | 1 |
| MC | Logan Hosp | 4 |
| MC | Mater Hlth | 1 |
| MC | Mayo | 110 |
| MC | Med Ctr Leeuwarden | 3 |
| MC | MedStar Hlth | 6 |
| MC | Metro South Hlth (Australia) | 1 |
| MC | Mt Area Health | 5 |
| MC | Myers JDC | 4 |
| MC | Natl Canc Ctr Sing | 13 |
| MC | Natl Hlthcare Grp (Singapore) | 1 |
| MC | North Shore | 5 |
| MC | Savannah Hlth Mission | 1 |
| MC | SE Priv Hosp | 1 |
| MC | STZ | 1 |
| MC | SW Hlthcare (Australia) | 1 |
| MC | Thasala Hosp | 1 |
| MC | Venizeleio Gen Hosp | 1 |
| MC | Vincent Van Gogh Inst Psychiat | 1 |
| NHS | Liverpool Womens | 6 |
| NHS | NHS | 70 |
| NHS | Oxfordshire | 1 |
| NR | Aga Khan Univ | 1 |
| NR | Ahvaz Jundishapur | 4 |
| NR | Albert Einstein Com | 3 |
| NR | All India Inst Med Sci | 1 |
| NR | Arabian Gulf | 1 |
| NR | Argosy Univ | 1 |
| NR | Ateneo Zamboanga Univ | 2 |
| NR | Austral Univ | 2 |
| NR | Baqiyatallah Univ | 1 |
| NR | Basel Inst Clin Epidemiol & Biostat | 2 |
| NR | Beijing Acad Educ | 1 |
| NR | Ben Gurion Univ Negev | 1 |
| NR | Blackburn Coll | 1 |
| NR | Brandon Univ | 2 |
| NR | Broadmoor | 1 |
| NR | Brock Univ | 2 |
| NR | Bucheon Univ | 1 |
| NR | Burrell Coll | 1 |
| NR | Busitema Univ | 1 |
| NR | Campbell Univ | 6 |
| NR | Campus Bio Med Univ | 3 |
| NR | Catholic Univ Daegu | 1 |
| NR | Catholic Univ Korea | 2 |
| NR | Cedarville Univ | 1 |
| NR | Cent Michigan Univ | 1 |
| NR | Chinese Acad | 5 |
| NR | Claremont | 2 |
| NR | Colorado Mt Coll | 1 |
| NR | CUNY | 1 |
| NR | CUSM | 1 |
| NR | Dankook Univ | 1 |
| NR | Dr. Alva Paramed Inst | 1 |
| NR | Duke-NUS | 10 |
| NR | Eastern Michigan | 1 |
| NR | EMESCAM | 1 |
| NR | European Univ | 1 |
| NR | FMMU | 8 |
| NR | Fujian Univ | 1 |
| NR | Gachon Univ | 1 |
| NR | Geisinger SOM | 1 |
| NR | Gulf Med Univ | 1 |
| NR | Hanze Univ | 2 |
| NR | Hatyai | 1 |
| NR | Heidelberg Univ | 3 |
| NR | Ho Chi Minh Cty Med Pharm Univ | 1 |
| NR | Hofstra | 5 |
| NR | Holmesglen Inst | 1 |
| NR | Icahn SOM | 6 |
| NR | Indiana Tech Univ | 1 |
| NR | Inje Univ | 1 |
| NR | Int Med Univ | 5 |
| NR | Ipswich Hosp | 1 |
| NR | Iuliu Hatieganu Univ | 1 |
| NR | Jeonju Univ | 1 |
| NR | Khyber Med Univ | 1 |
| NR | Kosin Univ | 1 |
| NR | Laureate Int Univ | 1 |
| NR | LECOM (Seton Hall) | 2 |
| NR | Loyola | 2 |
| NR | Macclesfield | 1 |
| NR | Martin Luther Univ | 2 |
| NR | MCPHS Univ | 1 |
| NR | Med Coll Wisc | 3 |
| NR | Meharry Med Coll | 8 |
| NR | Mercer Univ | 1 |
| NR | Molde Univ | 2 |
| NR | Mt Sinai | 6 |
| NR | Murdoch Univ | 6 |
| NR | NE Ohio Univ | 3 |
| NR | Nipissing Univ | 1 |
| NR | No Illinois Univ | 4 |
| NR | No Ontario SOM | 7 |
| NR | Ohio Univ | 1 |
| NR | Ontario Tech | 1 |
| NR | Open Univ (Amsterdam) | 1 |
| NR | Pacific Univ | 1 |
| NR | Paris Descartes Univ | 1 |
| NR | Peking Union Med Coll | 1 |
| NR | Peninsula | 6 |
| NR | PGIMER | 3 |
| NR | Princess Nora Univ | 1 |
| NR | Qassim Univ | 1 |
| NR | Queen Margaret Univ | 2 |
| NR | Quinnipiac Univ | 1 |
| NR | Red River Coll | 2 |
| NR | Republ Polytech | 1 |
| NR | Riphah Int Univ | 1 |
| NR | Ross Univ | 2 |
| NR | Royal Trop Ins | 2 |
| NR | Shifa Coll | 1 |
| NR | So Illinois | 10 |
| NR | So Methodist Univ | 1 |
| NR | St Georges | 1 |
| NR | St John Fisher Coll | 1 |
| NR | St Louis Univ | 10 |
| NR | Sunnybrook | 3 |
| NR | SUNY Buffalo | 6 |
| NR | SUNY Upstate | 4 |
| NR | SUNYIT | 4 |
| NR | Tele Univ | 1 |
| NR | Third Mil Med | 7 |
| NR | Thomas Jefferson | 10 |
| NR | Trinity Laban | 1 |
| NR | Uniformed Serv Univ | 43 |
| NR | Unitec | 1 |
| NR | Univ Akron | 1 |
| NR | Univ Ambrosiana | 1 |
| NR | Univ Appl Sci | 5 |
| NR | Univ Athens | 3 |
| NR | Univ Botswana | 6 |
| NR | Univ Buckingham | 2 |
| NR | Univ Calif San Diego | 2 |
| NR | Univ Calif San Francisco | 42 |
| NR | Univ Central Eng | 1 |
| NR | Univ Damman | 1 |
| NR | Univ Educ Freiburg | 1 |
| NR | Univ Fed Sao | 9 |
| NR | Univ Halle Wittenberg | 1 |
| NR | Univ Liege | 1 |
| NR | Univ London | 11 |
| NR | Univ Louisville | 5 |
| NR | Univ Lubeck | 1 |
| NR | Univ Lyon | 2 |
| NR | Univ Missouri | 3 |
| NR | Univ North Dakota | 1 |
| NR | Univ Notre Dame | 4 |
| NR | Univ Oklahoma | 1 |
| NR | Univ Osijek | 2 |
| NR | Univ Pretoria | 1 |
| NR | Univ Rotterdam | 1 |
| NR | Univ Rwanda | 1 |
| NR | Univ SE Norway | 4 |
| NR | Univ So Alabama | 1 |
| NR | Univ Texas Galveston | 1 |
| NR | Univ Texas Houston | 3 |
| NR | Univ Texas Med Branch | 2 |
| NR | Univ Texas SW Med Ctr | 4 |
| NR | Univ Vermont | 1 |
| NR | Univ Wales | 1 |
| NR | Univ Winchester | 5 |
| NR | Univ Witten | 6 |
| NR | Ursuline Coll | 1 |
| NR | USCS | 1 |
| NR | USF Odisseia | 1 |
| NR | Villanova Univ | 1 |
| NR | Virginia Commonwealth | 13 |
| NR | W China Hosp | 3 |
| NR | Waco Fam Med Res | 1 |
| NR | Walailak | 1 |
| NR | Walden Univ | 1 |
| NR | Wofford Coll | 1 |
| NR | Wright State | 5 |
| NR | Yeshiva Univ | 1 |
| NR | Yorkshire | 6 |
| ORG | AAMC | 2 |
| ORG | ABFM | 2 |
| ORG | ABIM | 9 |
| ORG | ABMS | 7 |
| ORG | Abor Aff No Dev Canada (AANDC) | 1 |
| ORG | ABP | 1 |
| ORG | ACGME | 9 |
| ORG | Aff Anesthes | 1 |
| ORG | AFMC | 1 |
| ORG | AKM STATS | 1 |
| ORG | AMA | 2 |
| ORG | AMEE | 1 |
| ORG | Amer Red Cross | 1 |
| ORG | APA | 1 |
| ORG | ASPPH | 1 |
| ORG | AXDEV | 4 |
| ORG | Belgian Hlth | 1 |
| ORG | Bloorview | 1 |
| ORG | BMJ | 1 |
| ORG | Booz Allen Hamilton | 1 |
| ORG | CAIPE | 2 |
| ORG | CAIR | 5 |
| ORG | Can Partnersh Canc | 1 |
| ORG | CASPolska | 4 |
| ORG | Catalan Hlth | 2 |
| ORG | CFPC | 1 |
| ORG | Chemplus | 1 |
| ORG | Childrens Natl Med Ctr (Wash) | 1 |
| ORG | China Exp & Cred Ins Corp | 1 |
| ORG | CHPCA | 1 |
| ORG | CIBERESP | 2 |
| ORG | CIHR | 1 |
| ORG | Clod Ensemble | 1 |
| ORG | Costa Rican Soc Secur Fund | 1 |
| ORG | CRO Aviano | 1 |
| ORG | Dept Hlth (Australia) | 1 |
| ORG | Dept of Veterans Affairs | 10 |
| ORG | Dept Publ Hlth (Florida) | 4 |
| ORG | Dutch Inspec Educ | 1 |
| ORG | Educ Commiss Foreign Med Grad | 4 |
| ORG | ENAULD Hlth Res | 1 |
| ORG | ETH | 1 |
| ORG | Fdn Adv Int | 2 |
| ORG | German Agcy Qual Med | 2 |
| ORG | GP Tng Val Coast | 1 |
| ORG | HealthPartners Inst | 3 |
| ORG | HESPER | 3 |
| ORG | Hlthy Afr Amer Fam | 1 |
| ORG | HOMER | 3 |
| ORG | Inst Lifestyle Med | 3 |
| ORG | Inst Work | 3 |
| ORG | Int Pov Red Ctr China | 1 |
| ORG | Intuit Surg | 2 |
| ORG | IUMSP (Switzerland) | 2 |
| ORG | Jump Sim Educ Ctr | 1 |
| ORG | Karolinska | 7 |
| ORG | London Deanery | 4 |
| ORG | London Learning | 1 |
| ORG | Maristan | 3 |
| ORG | Marshfield | 1 |
| ORG | Minist Hlth Accra | 1 |
| ORG | Minist Publ Hlth (Thailand) | 4 |
| ORG | Natl Ctr Poverty | 1 |
| ORG | Natl Fund Sci Res | 1 |
| ORG | NBME | 6 |
| ORG | NIH | 2 |
| ORG | NIH (Italy) | 1 |
| ORG | NIVEL | 3 |
| ORG | Norwegian Ctr E Hlth | 1 |
| ORG | NPS MedWise | 1 |
| ORG | Ortonville Area Svcs | 1 |
| ORG | Phil Museum Art | 3 |
| ORG | Rcsi | 8 |
| ORG | RealCME | 3 |
| ORG | Res & Expertise | 5 |
| ORG | Royal Australasian Coll Phys | 3 |
| ORG | Royal Australasian Coll Surg | 1 |
| ORG | Royal Coll Phys | 6 |
| ORG | Royal Coll Surg | 8 |
| ORG | RTI Int | 4 |
| ORG | SCC Assoc | 1 |
| ORG | SimPatiCo UK | 1 |
| ORG | SingHlth | 1 |
| ORG | Southern DHB | 1 |
| ORG | SRM Spokane | 1 |
| ORG | SSMG | 1 |
| ORG | St John God Clin Res Ctr | 1 |
| ORG | St Pau Biomed | 1 |
| ORG | THEnet | 1 |
| ORG | UCRH | 2 |
| ORG | US Dept Def | 1 |
| ORG | US FDA | 2 |
| ORG | Vict Inst Forens Med | 1 |
| ORG | WHO | 1 |
| ORG | WIcare | 1 |
| ORG | Work Psychol Grp | 9 |
| ORG | Yulius Acad | 1 |
| ORG | Zaniac | 1 |
| UNK | Rio de Janeiro (ESAM) | 1 |
| UNK | Unknown | 2 |
| #N/A | Not able to identify | 33 |
